# A Monte Carlo Estimation of the Narrow-Sense Heritability of COVID-19 Infection and Severity from AncestryDNA Survey Data

**DOI:** 10.1101/2022.05.23.22275364

**Authors:** Amelia J. Averitt, Deepika Sharma, Michael Cantor

## Abstract

Respiratory infectious diseases, such as COVID-19, demonstrate a host genetic component that contributes to interindividual differences of susceptibility and infection. At present, the relative effect of environmental and genetic factors of COVID-19 is unknown. This research presents a Monte Carlo (MC) estimation of the genetic narrow-sense heritability of COVID-19 infection and severity from AncestryDNA survey data. The results suggest a moderate genetic contribution to COVID-19 infection and a low genetic contribution for COVID-19 severity.

## Introduction

Many regard the emergence of COVID-19, caused by severe acute respiratory syndrome 2 (SARS-Cov-2), to be the greatest public health crisis of the past century. Effective management of infectious disease outbreaks, such as COVID-19, requires data-driven public health management. Often, public health management of infectious diseases is pathogen centered. This approach makes the assumption that the pathogen, itself, dictates the course of the disease. However, research has shown that genetics of the host may also contribute to infectious disease burden (Klebanov 2018). Respiratory infectious diseases, like COVID-19, also demonstrate a host genetic component that contributes to inter-individual differences (Patarčić et al. 2015). This contribution of the host’s genetics, in tandem with environmental factors, determines disease susceptibility and infection. Understanding the relative effect of environmental and genetic factors of respiratory infectious diseases may be helpful to guide public health intervention.

Genetic heritability summarizes the variation in observable characteristics – or phenotype – that is attributable to a variation in genetics rather than the environment or random effects. High genetic heritability indicates a strong phenotypic similarity between parents and offspring that is due to genetics, while low heritability indicates a low phenotypic similarity. A subset of genetic heritability is narrow-sense heritability (*h*^2^). *h*^2^ is the average proportion of total phenotypic variance that is due to additive genetic factors that are passed from parents to offspring. When discussing respiratory infectious diseases, knowledge of *h*^2^ can help inform public health interventions. In situations of uncertainty, as is the case with COVID-19, *h*^2^ estimates of disease susceptibility and severity may provide evidence of how the disease will, on average, affect families. This may enable clinicians to appropriately intervene on or monitor family groups and aid in allocation of medical resources.

Typically, heritability estimates, such as *h*^2^, are made via twin studies. In such studies, phenotypes of monozygotic (identical) twins are compared with phenotypes of dizygotic (fraternal) twins (Visscher, Hill, and Wray 2008). Though twin studies are robust to genetic and environmental confounders, they suffer from many limitations. They are inefficient, could result in an overestimation of heritability; and the study population may not be representative of the larger population, which could limit generalizability. Heritability estimates have also been made from the electronic health records, but often require lengthy patient recruitment, familial ascertainment, and phenotype identification (Polubriaginof et al. 2018).

## Methods

### The Method

This research presents a Monte Carlo (MC) estimation of the narrow-sense heritability of COVID-19 infection and severity from AncestryDNA survey data. This data is from a private collaboration with AncestryDNA and Regeneron Pharmaceuticals and was collected online from volunteer respondents. The survey was given to AncestryDNA customers (N210k) and aimed to assess each respondent’s COVID-19 infection status, exposure, risk factors, and symptoms. This data includes, if and which biological family members were infected with and hospitalized from COVID-19, which can be used for *h*^2^ estimation. Unlike traditional data for this task, the AncestryDNA survey data is borne from a large and heterogenous population, familial relationships are easily identified, and COVID-19 phenotypes are defined. Clinical data is preferred when making *h*^2^ estimates, but research demonstrates that self-reported data – such as survey data – yields concordance of heritability estimates from a sufficiently large sample (Macgregor et al. 2006).

In the circumstance when a parent is infected with or hospitalized from COVID-19, this survey’s responses fail to specify whether one or two parents were affected. This research uses an MC method to stochastically estimate the parent’s phenotypic variance using parameters that are grounded in real-world epidemiologic evidence. MC methods are a class of algorithms that rely on repeated sampling to make estimates that cannot be directly made. Over many sampling iterations, estimates derived from an MC procedure will theoretically converge with the true value. Our MC method is summarized in Algorithm 1. For this algorithm, if a respondent (proband) has one or more parents were infected or hospitalized, the count of afflicted parents is initialized to 1. Stochastic parameters for the simulation include the rate of two-parent households (*η*) (United States Census Bureau 2019); rate of household co-infection (*ρ*) (Li et al. 2020)(Grijalva et al. 2020)(Fung et al. 2021); and the rate of hospitalization among COVID infected individuals (*ψ*) (Garg et al. 2020). *h*^2^ is then estimated from the slope of a linear regression that models the simulated, mid-phenotypic value of the parents by the phenotypic value of the proband.

#### Algorithm 1

Pseudocode for Monte Carlo (MC) Estimation. Where, *N* is the number of iterations; *J* is number of observations in data; and *x*_*j*_ is number of parents in household for proband *j*.

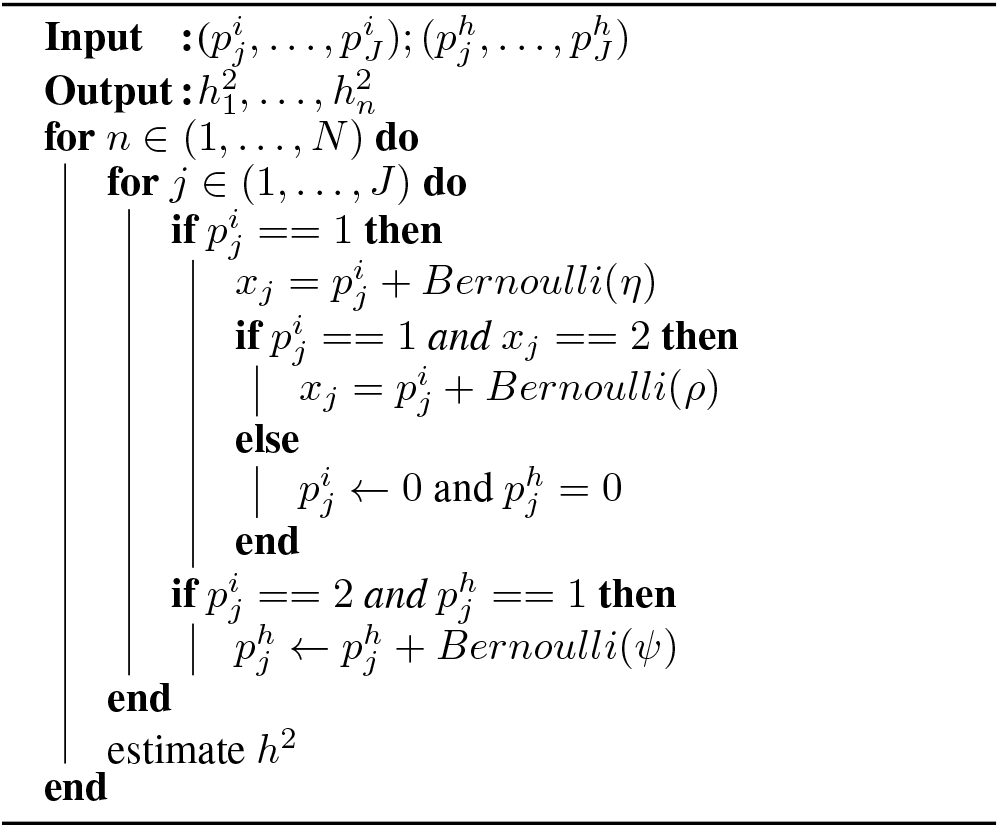

## Experimentation

This research presents (i) an application of the proposed methods to simulated data, and (ii) an application to AncestryDNA survey data to estimate *h*^2^ of COVID-19 infection 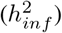 and severity 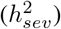

### Simulation

To evaluate the proposed method, we applied our estimation procedure to data in which the ground-truth *h*^2^ is known. Proband and parent phenotypes were simulated such that the ‘true’ *h*^2^ was hard-coded into the data. If 1 or 2 parents were affected, the data was then masked to only indicate if 1 or more was affected, which aligns with the uncertainty in the AncestryDNA data. The MC algorithm was applied to this masked, simulated data and an *h*^2^ estimate was made. This experimental set-up was repeated 1000 times using varying ‘true’ *h*^2^ values of 0.1, 0.3 and 0.5.

## Results

When applied to simulated data, in which groundtruth *h*^2^ is known, the method is able to recover *h*^2^ with high accuracy (Table 1). Presuming that the parameters are true, this indicates that the proposed method can recover *h*^2^ with minimal bias.

**Table 1:** Mean MC estimates and 95‥ confidence intervals of *h*^2^ when applied to masked, simulated data with target *h*^2^s of 0.1, 0.3, and 0.5.

| Target $h^2$ | MC $h^2$ estimate |
| --- | --- |
| **0.1** | 0.103 [0.079, 0.124] |
| **0.3** | 0.323 [0.302, 0.342] |
| **0.5** | 0.479 [0.469, 0.494] |

### Application to AncestryDNA Survey Data

To estimate *h*^2^ from the AncestryDNA data, proband phenotypes for COVID-19 infection and severity were binarized. Cases of proband COVID-19 infection (2.24%) were identified as respondents who reported any of the following (i) a swab-test for COVID-19 that was positive; (ii) no swab-test for COVID-19, but respondent had flu-like symptoms after February 2020; or (iii) an antibody test for COVID-19 that was positive. From the population of infection cases, severe COVID-19 (0.25%) probands were defined as respondents who reported any of the following (i) hospitalization due to COVID-19; (ii) hospitalization in the ICU with oxygen; or (iii) hospitalization in the ICU with a ventilator. Cases were assigned a phenotype of 1. In the absence of case requirements for either phenotype, the proband phenotype was 0. Parents of a proband that did not report that their “Parent(s)” were infected or hospitalized due to COVID-19 were assigned a phenotype of 0 for both outcomes. In the event that a proband reported that “Parent(s)” were infected or hospitalized due to COVID-19, this data was passed into the MC simulation to stochastically model the parent phenotype; if 1 or 2 parents were infected (*p*^*i*^) and hospitalized (*p*^*h*^). Models for both phenotypes of interest, were adjusted for the age, gender, and ever-smoking status of the proband. Given the high variability in reported household co-infection, *ρ*, the MC simulation was repeated using three reported statistics to inform this parameter (Li et al. 2020)(Grijalva et al. 2020)(Fung et al. 2021).

### Results

The results of the application to AncestryDNA survey data indicate that *h*^2^ of COVID-19 infection susceptibility ranges from 0.1554 to 0.1833. 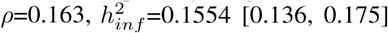 when 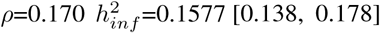; and when 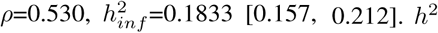. *h*^2^ of COVID-19 severity ranges from 0.0734 to 0.0751. When 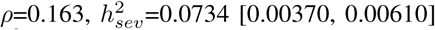 when 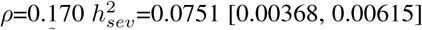and when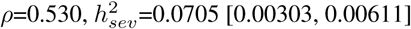These results sug-gest a moderate genetic contribution to COVID-19 infection and a low genetic contribution for COVID-19 severity (Figure 1). For context, *h*^2^ of height is 0.8 (Macgregor et al. 2006).

**Figure 1:**
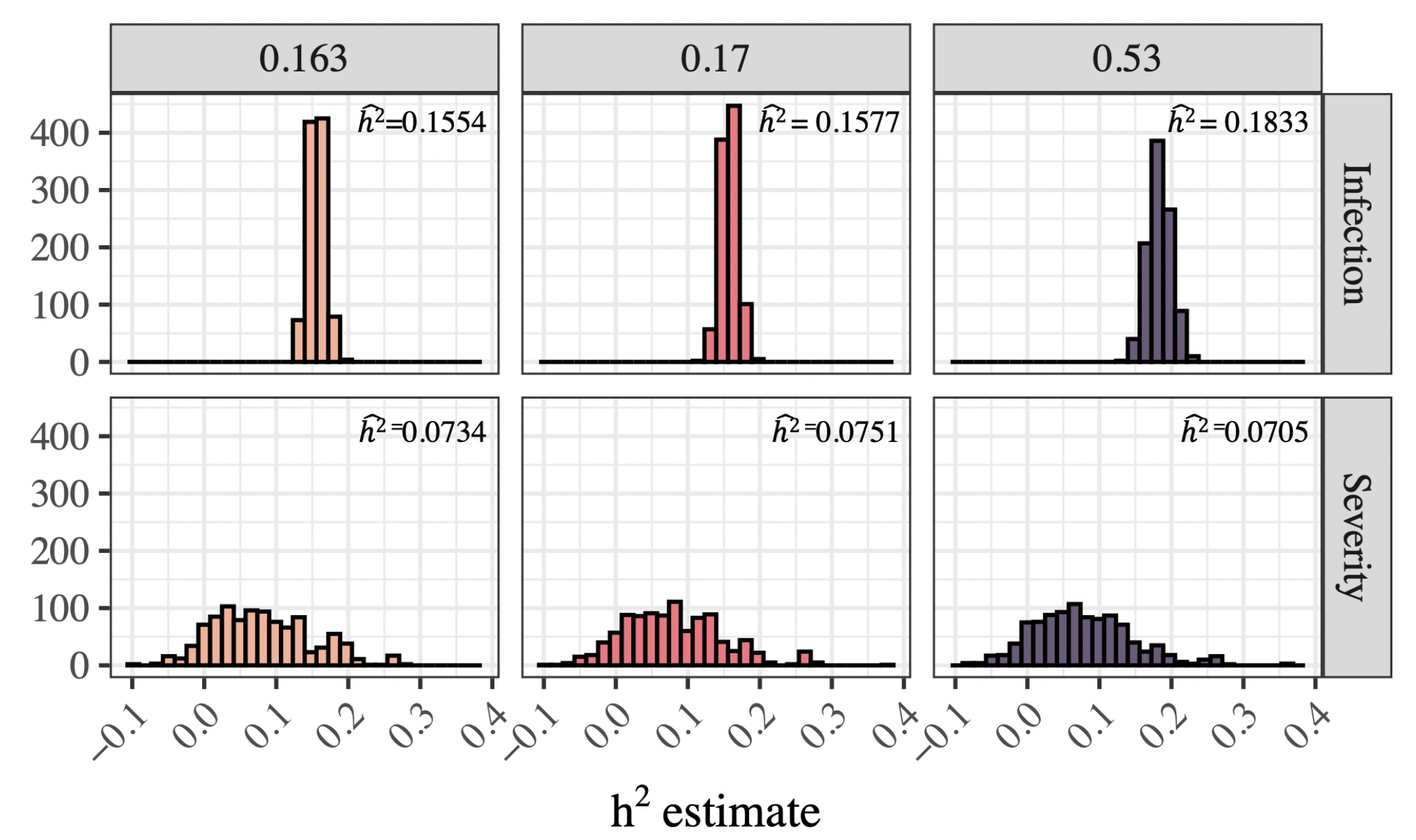
MC estimates of *h*^2^ from AncestryDNA data, with varying parameters of household co-infection – 0.163 (Li et al. 2020), 0.170 (Grijalva et al. 2020), and 0.530 (Fung et al. 2021).

## Discussion

This research provides evidence that variability in infection by SARS-Cov-2 is, in part, explained by inherited genetic factors. Clinicians and public health professionals should be mindful of inherited disease susceptibility and monitor biological family members of the infected. These estimates may also provide context for genetic contribution to infection severity and hospitalization for disease variants and other pathogens. However, this approach does not account for recessive effects or gene-gene interaction and is reliant upon the estimated parameters. furthermore, this research and abstract was completed in March 2020. COVID research, including our understanding of genetic mechanisms of disease and epidemiologic parameters, have matured since this time.

### Ethical Statement

All data for this research project was from individuals who provided prior informed consent to AncestryDNA, as reviewed and approved by our external institutional review board, Advarra (formerly Quorum). All data were de-identified before use.

## Data Availability

Due to privacy and ethical concerns, supporting data cannot be made openly available.

## References

[Fung et al. 2021] Fung, H. F.; Martinez, L.; Alarid-Escudero, F.; Salomon, J. A.; Studdert, D. M.; Andrews, J. R.; Goldhaber-Fiebert, J. D.; Chin, E. T.; Claypool, A. L.; Fernandez, M.; Gracia, V.; Luviano, A.; Rosales, R. I. M.; Reitsma, M.; Ryckman, T.; and Ryckman, T. 2021. The Household Secondary Attack Rate of Severe Acute Respiratory Syndrome Coronavirus 2 (SARS-CoV-2): A Rapid Review. Clinical Infectious Diseases 73(Supplement<sub>2</sub>) : S138––S145.

[Garg et al. 2020] Garg, S.; Kim, L.; Whitaker, M.; O’Halloran, A.; Cummings, C.; Holstein, R.; Prill, M.; Chai, S. J.; Kirley, P. D.; Alden, N. B.; Kawasaki, B.; Yousey-Hindes, K.; Niccolai, L.; Anderson, E. J.; Openo, K. P.; Weigel, A.; Monroe, M. L.; Ryan, P.; Henderson, J.; Kim, S.; Como-Sabetti, K.; Lynfield, R.; Sosin, D.; Torres, S.; Muse, A.; Bennett, N. M.; Billing, L.; Sutton, M.; West, N.; Schaffner, W.; Talbot, H. K.; Aquino, C.; George, A.; Budd, A.; Brammer, L.; Langley, G.; Hall, A. J.; and Fry, A. 2020. Hospitalization Rates and Characteristics of Patients Hospitalized with Laboratory-Confirmed Coronavirus Disease 2019 — COVID-NET, 14 States, March 1–30, 2020. MMWR. Morbidity and Mortality Weekly Report 69(15):458–464.

[Grijalva et al. 2020] Grijalva, C. G.; Rolfes, M. A.; Zhu, Y.; McLean, H. Q.; Hanson, K. E.; Belongia, E. A.; Halasa, N. B.; Kim, A.; Reed, C.; Fry, A. M.; and Talbot, H. K. 2020. Transmission of SARS-COV-2 Infections in Households — Tennessee and Wisconsin, April–September 2020. MMWR. Morbidity and Mortality Weekly Report 69(44).

[Klebanov 2018] Klebanov, N. 2018. Genetic Predisposition to Infectious Disease. Cureus 10(8).

[Li et al. 2020] Li, W.; Zhang, B.; Lu, J.; Liu, S.; Chang, Z.; Peng, C.; Liu, X.; Zhang, P.; Ling, Y.; Tao, K.; and Chen, J. 2020. Characteristics of Household Transmission of COVID-19. Clinical infectious diseases : an official publication of the Infectious Diseases Society of America 71(8):1943–1946.

[Macgregor et al. 2006] Macgregor, S.; Cornes, B. K.; Martin, N. G.; and Visscher, P. M. 2006. Bias, precision and heritability of self-reported and clinically measured height in Australian twins. Human genetics 120(4):571–80.

[Patarčić et al. 2015] Patarčić, I.; Gelemanović, A.; Kirin, M.; Kolčić, I.; Theodoratou, E.; Baillie, K. J.; de Jong, M. D.; Rudan, I.; Campbell, H.; and Polašek, O. 2015. The role of host genetic factors in respiratory tract infectious diseases: systematic review, meta-analyses and field synopsis. Scientific Reports 5(1):16119.

[Polubriaginof et al. 2018] Polubriaginof, F. C. G.; Vanguri, R.; Quinnies, K.; Belbin, G. M.; Yahi, A.; Salmasian, H.; Lorberbaum, T.; Nwankwo, V.; Li, L.; Shervey, M. M.; Glowe, P.; Ionita-Laza, I.; Simmerling, M.; Hripcsak, G.; Bakken, S.; Goldstein, D.; Kiryluk, K.; Kenny, E. E.; Dudley, J.; Vawdrey, D. K.; and Tatonetti, N. P. 2018. Disease Heritability Inferred from Familial Relationships Reported in Medical Records. Cell 173(7):1692–1704.e11.

[United States Census Bureau 2019] United States Census Bureau. 2019. Families Living Arrangements.

[Visscher, Hill, and Wray 2008] Visscher, P. M.; Hill, W. G.; and Wray, N. R. 2008. Heritability in the genomics era — concepts and misconceptions. Nature Reviews Genetics 9(4):255–266.

